# A statistical forecast of LOW mortality (< 400,000 deaths) due to COVID-19, for the whole WORLD

**DOI:** 10.1101/2020.04.26.20074377

**Authors:** Cesar A. Barbero

**Affiliations:** Departamento de Química, Facultad de Ciencias Exactas, Fisicoquímicas y Naturales, Universidad Nacional de Rio Cuarto; Instituto de Investigaciones en Tecnologías Energéticas y Materiales Avanzados

## Abstract

**OBJECTIVE:** To forecast the death toll of COVID-19 in the whole world by fitting the time series of reported deaths with a parametric equation (integrated Gaussian equation) related to Farr’s law.

**DATA:** The time series of cumulative deaths due to COVID-19 produced by John Hopkins University and stored in a github repository.

**RESULTS:** The projected total death toll will be **261680** (392520 – 183176) which represents the 0.003 % of world population. This number amounts to 0.054 deaths per 1000, while the mean in the world (all causes) is 7.7. The daily peak of deaths (**7270** (+/-500)) happened the 15 (+/- 3) of April, meaning that we are in descending curve of the pandemic. The outbreak will end completely the 23^th^ (+/-3) of June. However, already on 9^th^ (+/- 3) of May, 2 **σ** (95.45%) of the deaths will have be occured. The projected death toll is much lower (5-10 times) than those forecasted by the Imperial College Group (ICG) even considering the best scenario of total suppression of virus transmission. Using actual mortality rates it is possible to back calculate which number of infected individuals would produce such mortality. The death toll arises from a number of infected individuals between **53 (worst case) and 3**.**3 million**. The calculated number of infected individuals is significantly lower than that calculated by ICG (227.5 millions) with suppression.

## Introduction

The COVID-19 disease seems to be the most important crisis since the influenza pandemic in 1918-1919. The disease is caused by the coronavirus SARS-Cov-2 virus. Since there is no vaccine for such novel virus and seems that there is no accepted antiviral therapy against it, the only way to avoid the disease is avoid its propagation. However, the lack of vaccine and antiviral therapy is usually the case with common cold and new strain of influenza (like H1N1 in 2009.[1]) The unusual circumstance here is that COVID-19 seems to be highly contagious (R0 = 2-3) compared with influenza (R0= 1.25).[2] The mortality coefficient (Rm= deaths/infected) could be quite high (4-8 %) but depends heavily on detecting all infected individuals, even those asymptomatic.[3] The The World Health Organization (WHO) declares COVID-19 a pandemic 13 of March.[4] At present (20^th^ of April), the number of deaths is of more than 160.000 [3] COVID-19 pandemic is at early stages and the number of cases and deaths could grow considerably. The capability of forecasting relevant parameters (deaths, critical or grave cases) is critical.

Plenty of models have been set-up to model the number of cases and deaths of COVID-19. Most rely on SIR (Susceptible-Infected-Removed) simulation using systems of differential equations.[5] A well-known calibrated SRI simulation was made by the Imperial College Group,[5] whom inform the WHO and guide policy in the world. They predict 7.0 billion infections (89 % of world population) will be infected and this would cause 40 million deaths (Rm = 0.57%) if no containment action is set in place.[6] The number decrease to 20 million deaths if mitigation (equivalent to those implemented during H1N1 pandemic) and to 9.3 million deaths if the extreme containment (“suppression”) is applied after a threshold: 1.6 deaths per 100,000 population per week) (high threshold,HT) has been reached. The best case scenario implies application of suppression before the number of deaths has reached 0.2 deaths per 100,000 population per week (low threshold, LT). In that case, 1.3 million will die of COVID-19.[6] It should be mentioned that, while the LT for the world was reached on 28^th^ of March, the HT has not been reached yet. Since, according to WHO, 650.000 died every year of seasonal flu,[7] anything short of extreme containment will produce avoidable excess deaths. However, as we will see below, we forecast a total number of deaths several times (5 to 6 times) lower than the best case scenario produced by ICG.

The Institute for Health Metrics and Evaluation (IHME) of Washington University,[18] uses a fitting method to obtain a parametric equation which allows forecasting the number of deaths during time using the data of the ongoing outbreak. The equation that describes the number of deaths per day is the Gaussian distribution equation. The total number of deaths per day is the integral form of the equation. The equation represents a bell curve which is the shape found out empirically by Farr (Farr’s law) which have found to fit most of epidemic outbreaks of diseases. While IMHE models several countries of the world, it does not apply the methodology to the whole world. For informing health and economic policy such information is necessary.

Therefore, programs were set to fit the integral of the Gaussian equation. The fitting and forecasting capabilities of the programs were tested with advanced cases (in the northern hemisphere) where can be compared with IHME data. Then, they were applied to Argentina and southern hemisphere countries. [8] The forecast for countries in the southern hemisphere show relatively low excess deaths. The calculation of different scenarios for infection rates also shows a relatively low infection rates.

The repository of cumulative numbers (time series) of deaths in github,[9] supported by the John Hopkins University is a large data file with comma separated values (csv). Simply importing in a spreadsheet program (like LibreOffice) allows calculating relevant numbers (like the number of deaths per week per 100,000 shown above). Moreover, it also allows calculating the time series of the cumulative number of deaths for the whole world. The pandemic nature of the COVID-19 assures that the region of the outbreak is the whole world. Obviously, the shutdown of international travel change the situation but it is equivalent to the shutdown of internal travel in a big country like China (which also represents the 17% of the world population) which is one of the mitigation measures. Applying the procedure to the world predicts that less than 400,000 deaths will be caused by COVID-19.

### Experimental

The program was described in a previous publication, [8] It was developed in Python 3.76 in the IDE Spyder launched by Anaconda Navigator on a laptop (Dell Latitude 3460 laptop. The Gaussian equation (Farr’s law) was symbolically integrated with Euler by adding integration constant. Using functions in Python ([10]) written in a Python program (available at [11]).

The test suite used was the data from Italy, Spain and Iran that contains more than 2/3 of the peak (deaths per day). The program predicts the same parameters using 2/3 of the data from the country or 1/3, showing that it is able to adequately predict the evolution of deaths over time. The data was produced by John Hopkins University and managed in a github repository [9]. In that way, the database is public and its quality is independently verified.

A special program: “Graphical fit” was used to obtain roughly correct parameters (a,b,c, norm) to be used as seed and as central points for the constraint of the curve fitting routine. By trial and error, guided by the understanding of each parameter on the shape of the curve, the simulated data is graphed along the experimental data. When a reasonable fit is obtained, the parameters are copied in the curve fitting program as seed. The constraints of the curve fit are set around those values. The curve fitting program is run until the parameters are different to the constraints. The numerical and graphical output is directly written in the manuscript, without any data manipulation. The results of the fit (numerical data and plots) for each country are provided in the supplementary information and in github.

## Results and discussion

### Assessing relevant parameters of the world

The fitting of the reported cumulative deaths data is shown in Figure 1.

**Figure 1.**
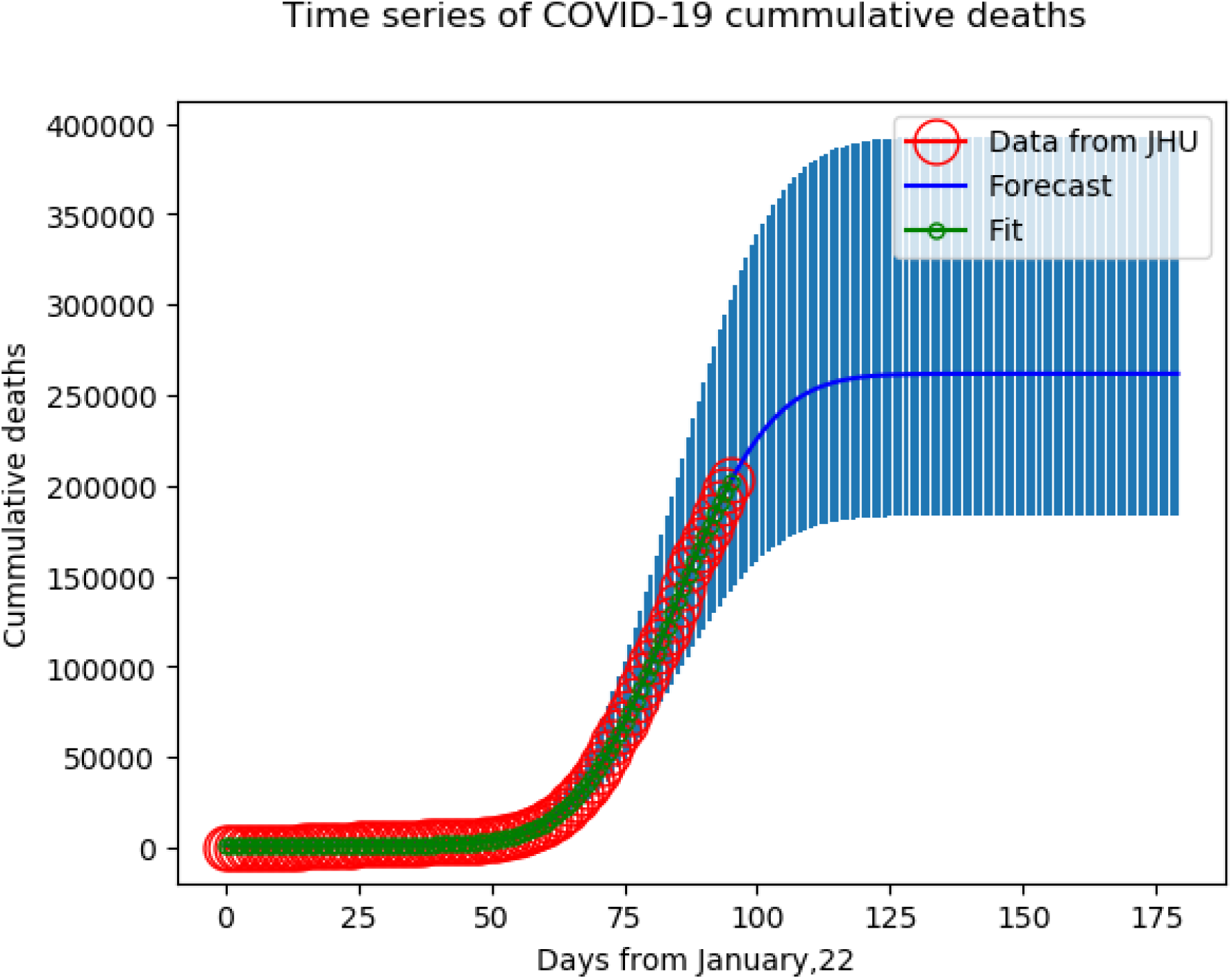
Plot of the time (days) series of cumulative deaths by COVID-19 in the WORLD (red circles). The green squares represent the fitted data while the blue line shows the integral of the gaussian equation with the parameters produced by the fit. The dashed blue vertical lines represent the uncertainty of the predicted data..

The fitting is quite good and allow forecasting the relevant parameters. Total deaths = **261680** (392520 - 183176). The total number of deaths amounts to the 20-30% of the number predicted for the best case scenario (75% suppression) by the ICG.[6] It also amounts to 0.0034 % of the world population. It is also smaller than deaths due to seasonal flu.[7]

Other important parameters are the day of maximum death since it is related to the day of maximum usage of the health system. To find out, the Gaussian equation is plotted along with the data of deaths per day. (Figure 2).

**Figure 2.**
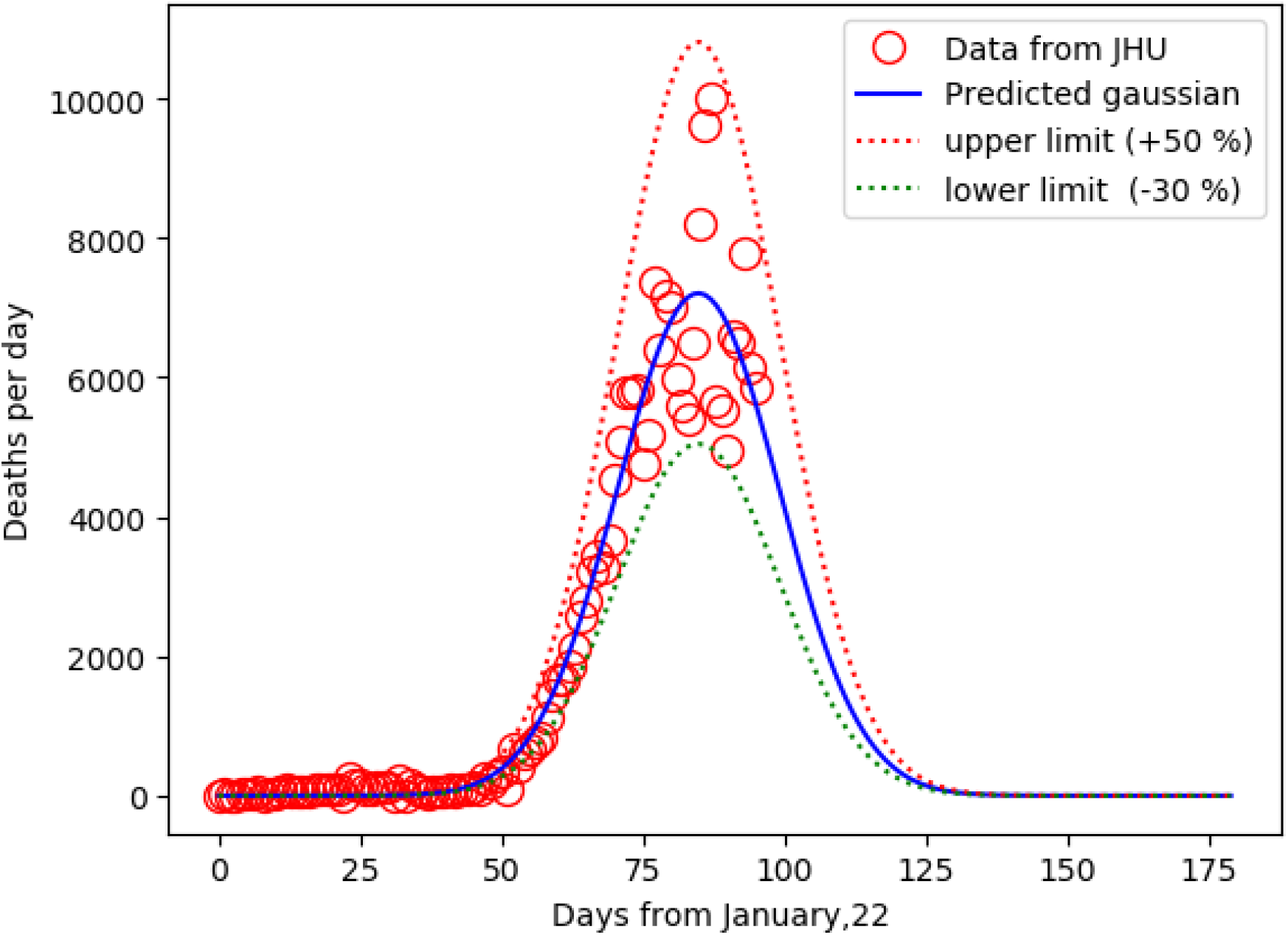
Graph of the deaths per day (red circles) along with the parametric Gaussian equation plot (blue line). calculated using the parameters obtained from the fit The red and green dashed lines represent the upper and lower limit of uncertainty.

In the case of the World, the maximum, **7209** (+/-500), deaths happened the 15 (+/-3) of April. Note that the reported data of deaths per day show a large scatter due to the difficult reporting deaths in the whole world. The large scatter makes difficult to fit directly the Gaussian equation but 95% of the actual point occurs inside the uncertainty range (50%).

The fact that the peak number of deaths occurs around 15^th^ of April allows a simple calculation of the total number of deaths. Since the Gaussian equation is a symmetrical curve, the peak defines two equal areas. The area of the curve is the integral, representing the number of cases. Therefore, the total number of cases should be equal to twice the number of deaths on 15^th^ of April. According to the JHU data, the number of deaths was 125983, therefore the number of deaths at the end of the pandemic outbreak should be: 251966, which is slightly below the number calculated by the program (261680).

Additionally, the equation allows calculating the end of the outbreak which is the 23^th^ (+/-3) of June. However, already the 9^th^ (+/-3) of May, 95.45% (2 σ) of the deaths have occurred. It is reasonable to think that at that date, the outbreak is over.

As it can be seen, the PIGE predicts a relatively low number of deaths. Not all the countries of the world decide to apply a full lockdown (and others have difficulties to enforce total lockdown) but the outcomes are everywhere dramatically better than the IGC estimation. Therefore it is reasonable to assume that the IGC model,[6] overestimate the number of deaths.

The parametric equation allows detecting the maximum of deaths per day peak. The world has already passed the peak meaning that the outbreak is subsiding. Since the number of deaths have to be related with the number of new infections (with a delay), it also means that number of new cases is decreasing. In any epidemiological model, this means that the number of infections caused by an infected individual is decreasing in time. This could be to the fact that the number of susceptible contacts is decreasing (because all are already infected or recovered) or that the number of contacts is decreasing with time. A way to calculate the number of persons infected involves dividing by the rate of mortality (Rm = deaths/infected). However, Rm is not easily known because it would require knowing exactly the number of persons infected in all the countries involved, and such number depends on the testing capabilities and testing protocol. In Argentina, only individuals who show symptoms are tested, therefore asymptomatic infected individuals are not counted. Since different Rm values have been reported, the Rm reported by South Korea (0.5 %) was used to produce the high limit number and that of Italy (8 %) to calculate the lower limit. Using these assumptions, a worst case scenario of 53,050,000 (+/-10000) individuals should be infected.

The number is relatively low since the population of the world is of more than 7,300M (represents the 0,007 %). The report by ICG consider an infection rate of 7 billion (89 %) for the unmitigated response and 227,500,000 (0.03 %) for the total suppression. The latter is larger (ca. 4 times) than our forecasting.

## Conclusions

The fitting with a parametric integrated Gaussian equation (PIGE) seems to be a good method to obtain parameters of both descriptive and predictive value for the COVID-19. Using open software libraries in Python (3.7) it is possible to produce programs which fit the experimental data (reported cumulative deaths). The application to the reported data for the whole world allows forecasting a relatively small number of deaths for the whole world. It also allows calculating: i) the day of the peak and the day of the end of the outbreak.

Using the parameterized death data and the mortality ratio (Rm), it is possible to calculate the number of COVID-19 cases, assuming two scenarios: a low Rm (South Korea) and a high Rm (Italy). These assumptions translate into a worst case scenario and best case scenario, respectively, for the number of infected individuals. These data are also much smaller than those predicted by the ICG.

## Data Availability

A set of document files with the results, datasets and the programs are available at https://github.com/cesarbarbero/programas-para-predecir-COVID-19/).

## Acknowlegements

This work was funded by Universidad Nacional de Rio Cuarto (UNRC, Argentina) and the Consejo Nacional de Investigaciones Cientificas y Tecnicas (CONICET, Argentina) whom paid my salary while I am socially isolated at home. Helpful virtual discussions with J. Balach and E. Quinteros about the program are also discussed. The laptop used was purchased with funds from a Secretaria de Politicas Universitarias – Minsterio de Educacion (Argentina). The author appreciates the use of open source programs (Python, Anaconda, Spyder, Matplotlib, Scipy, Numpy, Euler Math Toolbox, Vseuz graphics software). Finally, special thanks to Dr. L. Sereno whom teach me how to visually find out good seed/constraints for nonlinear fitting of complex equations.

## References

[1] https://www.who.int/csr/disease/swineflu/en/

[2] https://www.who.int/news-room/q-a-detail/q-a-similarities-and-differences-covid-19-and-influenza

[3] Covid-19: identifying and isolating asymptomatic people helped eliminate virus in Italian village,Day, Michael, BMJ, 2020/03/23, 10.1136/bmj.m1165

[4] http://www.euro.who.int/en/health-topics/health-emergencies/coronavirus-covid-19/news/news/2020/3/who-announces-covid-19-outbreak-a-pandemic.

[6] Patrick GT Walker, Charles Whittaker, Oliver Watson et al. The Global Impact of COVID-19 and Strategies for Mitigation and Suppression. Imperial College London (26-03-2020), doi: https://doi.org/10.25561/77735.

[7] https://www.who.int/news-room/detail/14-12-2017-up-to-650-000-people-die-of-respiratory-diseases-linked-to-seasonal-flu-each-year

[8] medRxiv (MS ID#: MEDRXIV/2020/072488 MS TITLE: A statistical forecast of LOW mortality and morbidity due to COVID-19, in ARGENTINA and other Southern Hemisphere countries.

[9] https://github.com/CSSEGISandData/COVID-19/blob/master/csse_covid_19_data/csse_covid_19_time_series/time_series_covid19_deaths_global.csv#L17

[10] scipy.optimize, https://www.scipy.org/

[11] https://github.com/cesarbarbero/programas-para-predecir-COVID-19/

